# A Deep Learning Model for Brain Age Prediction Using Minimally Pre-processed T1w-images as Input

**DOI:** 10.1101/2022.09.06.22279594

**Authors:** Caroline Dartora, Anna Marseglia, Gustav Mårtensson, Gull Rukh, Junhua Dang, J-Sebastian Muehlboeck, Lars-Olof Wahlund, Rodrigo Moreno, José Barroso, Daniel Ferreira, Helgi B. Schiöth, Eric Westman, Alzheimer’s Disease Neuroimaging Initiative, Australian Imaging Biomarkers and Lifestyle flagship study of ageing, Japanese Alzheimer’s Disease Neuroimaging Initiative, AddNeuroMed consortium

**Author notes:** Corresponding authors: Caroline Dartora and Eric Westman Karolinska Institutet Department of Neurobiology, Care Sciences and Society (NVS) Division of Clinical Geriatrics Neo floor 7 | SE 141 83 Huddinge Sweden. Data used in the preparation of this article were obtained from the Alzheimer’s Disease Neuroimaging Initiative (ADNI) database (adni.loni.usc.edu). As such, the investigators within the ADNI contributed to the design and implementation of ADNI and/or provided data but did not participate in the analysis or writing of this report. A complete listing of ADNI investigators can be found at: http://adni.loni.usc.edu/wp-content/uploads/how_to_apply/ADNI_Acknowledgement_List.pdf. Data used in the preparation of this article was obtained from the Australian Imaging Biomarkers and Lifestyle flagship study of aging (AIBL) funded by the Commonwealth Scientific and Industrial Research Organisation (CSIRO), which was made available at the ADNI database (www.loni.usc.edu/ADNI). The AIBL researchers contributed data but did not participate in the analysis or writing of this report. AIBL researchers are listed at https://aibl.csiro.au/. Data used in the preparation of this article were obtained from the Japanese Alzheimer’s Disease Neuroimaging Initiative (J-ADNI) database deposited in the National Bioscience Database Center Human Database, Japan (Research ID: hum0043.v1, 2016). As such, the investigators within J-ADNI contributed to the design and implementation of J-ADNI and/or provided data but did not participate in the analysis or writing of this report. A complete listing of J-ADNI investigators can be found at: https://humandbs.biosciencedbc.jp/en/hum0043-j-adni-authors.

## Abstract

In the last few years, several models trying to calculate the biological brain age have been proposed based on structural magnetic resonance imaging scans (T1-weighted MRIs, T1w) using multivariate methods and artificial intelligence. We developed and validated a convolutional neural network (CNN)-based biological brain age prediction model that uses only one T1w MRI pre-processing step to simplify implementation and increase accessibility in research settings. Our model only requires rigid image registration to the MNI space, which is an advantage compared to previous methods that require more pre-processing steps, such as feature extraction. We used a multicohort dataset of cognitively healthy individuals (age range = 32.0 – 95.7 yrs.) comprising 17296 MRIs for training and evaluation. We compared our model using hold-out (CNN1) and cross-validation (CNN2-4) approaches. To verify generalizability, we used two external datasets with different population and MRI scan characteristics to evaluate the model. To demonstrate its usability, we included the external dataset’s images in the cross-validation training (CNN3). To ensure that our model used only the brain signal on the image, we also predicted brain age using skull-stripped images (CNN4). The trained models achieved a mean absolute error of 2.99, 2.67, 2.67, and 3.08 yrs. for the CNN1-4, respectively. The model’s performance in the external dataset was in the typical range of mean absolute error (MAE) found in the literature for testing sets. Adding the external dataset to the training set (CNN3), overall, MAE is unaffected, but individual cohort MAE improves (2.25 to 5.63 years). Salience maps of predictions reveal that periventricular, temporal, and insular regions are the most important for age prediction. We provide indicators for using biological (predicted) brain age as a metric for age correction in neuroimaging studies as an alternative to the traditional chronological age. In conclusion, using different approaches, our CNN-based model showed good performance using only one T1w brain MRI pre-processing step. The proposed CNN model is made publicly available for the research community to be easily implemented and used to study aging and age-related disorders.

## 1 INTRODUCTION

In recent years, the concept of an individual’s biological age—which can differ from the person’s chronological age—has sparked great interest in the medical research community, as ageing is a significant risk factor for several age-related health conditions and mortality. There is also substantial heterogeneity in health outcomes among individuals of the same chronological age (Jylhävä et al., 2017). During the past decades, the research highlighted that the biological aging process varies between people because of the complex interplay between genetic and environmental factors, such as lifestyle behaviours (Cole et al., 2019, 2017; Fratiglioni et al., 2020). Chronological age is the primary risk factor for most neurodegenerative disorders, including Alzheimer’s disease (AD) and vascular dementia (Hou et al., 2019). Like other age-related health conditions, also in the dementia field, there is significant heterogeneity in the manifestation of the symptoms as well as underlying brain pathology between people of the same chronological age (Ferreira et al., 2020). Therefore, quantifying the biological age could be a better metric than the traditional chronological age to identify individuals at risk of developing age-related diseases (Cole et al., 2019; Tian et al., 2023).

Parallel advancements in neuroscience and computational science have enabled researchers to develop novel algorithms to determine the biological age of the brain. A biological marker of brain age will enable us to adjust neuroimaging studies for the person’s biological age instead of chronological age, minimising anatomical and functional heterogeneities present in groups of healthy individuals. Another advantage is that this could lead to a deeper understanding of pathological ageing mechanisms, which can culminate in dementia. Dementia is a multifactorial syndrome in which decades of accumulating neuropathology precedes clinical manifestation (Jack et al., 2010). The loss of neurons and synapses during the preclinical and prodromal stages can lead to brain atrophy and, therefore, to “older-looking” brains (when biological brain age, i.e. predicted age, is higher than chronological age) (Bashyam et al., 2020; Cole, 2020; Cole and Franke, 2017; Elliott et al., 2021; Franke and Gaser, 2012; Glorioso et al., 2019; Hwang et al., 2020). In contrast, some individuals will show higher chronological age than the biological brain age, thus showing a “younger-looking” brain, which could reflect relatively preserved brain structures (e.g., brain maintenance and/or cognitive reserve) (Cole et al., 2019; Stern et al., 2020). With the unprecedented growth of the elderly population worldwide, the expected increase in dementia cases (WHO guidelines, 2019), and while waiting for effective pharmacological treatments against dementia, a biological marker of brain age could play a key role in dementia prevention (Brusini et al., 2022).

In the past few years, several brain age models have been developed using different methods to estimate the age of the brain (Baecker et al., 2021a; Bocancea et al., 2021). These prior studies used machine (Cole et al., 2017; Franke et al., 2010; Franke and Gaser, 2012; Hwang et al., 2021) and deep learning (Bintsi et al., 2020; Cole et al., 2017; Gupta et al., 2021; Jonsson et al., 2019; Kolbeinsson et al., 2019; P. Lam et al., 2020a; Liang et al., 2019; Niu et al., 2020) approaches, achieving good performance in terms of mean absolute error (MAE) – between 2 and 6 years. However, the model type and the input choice varied across these studies that used pre-processed magnetic resonance imaging (MRI) data (T1-weighted, T1w), going through normalisation, corrections, segmentation steps, or even image feature extraction. Such a chain of steps is challenging to implement in research and, in the long term, in clinical settings due to time- and resource-consuming constraints.

Typically, a model is trained on neuroimaging data of healthy individuals from one or multiple cohorts covering a large age span. Developing a model for predicting “biological brain age” depends on the choice of training data when defining healthy aging. The “ideal” dataset would include: (1) detailed information and clinical data of study participants in order to be as comprehensive as possible with the selection criteria; (2) a large set of images, which are required to train a CNN model (Sajedi and Pardakhti, 2019); (3) participants with a diverse demographical background and a large, preferably uniform, age distribution to apply the developed model in more datasets (i.e., increase generalizability); (4) images acquired with a wide range of MRI scanners and protocols to improve generalisation to new unseen data of the model (Mårtensson et al., 2020), and (5) longitudinal data to ensure that the model does not predict a lower age at a later time point. Although initiatives to gather large-scale population-based datasets are ongoing (e.g., UK Biobank), to our knowledge, currently, no existing cohort possesses all the five characteristics listed above.

In this study, we aimed to develop and validate a CNN model, based on brain images, that uses only one pre-processing step (i.e., rigid registration of T1w MRIs to the Montreal Neurological Institute – MNI – template space) for brain age prediction. This minimal pre-processing feature has the advantage and strength of simplifying the model’s implementation and increasing accessibility in research settings. When publicly available, the model can be quickly used for any T1w MRI scan without time- and resource-consuming pre-processing steps. To evaluate our model, we used a large dataset of cognitively healthy individuals from six cohorts to address the “ideal” dataset criteria. The CNN model was compared using hold-out and cross-validation approaches. To verify the model’s generalizability, we tested the abovementioned approaches using two external datasets containing different scanners and demographic characteristics from the training set. Furthermore, we included the two cohorts used as external datasets in the cross-validation loop to verify the model’s usability with different cohorts. Finally, we employed the cross-validated model to predict brain age in skull-stripped images to ensure our model accurately predicted based on the brain image signal. We then evaluated the model’s performance using two external datasets.

## 2 METHODS

### 2.1 STUDY POPULATION

For this study, we included 17296 T1w MRIs from 15289 (1176 are 1.5T and 16120 are 3T) cognitively healthy participants from six cohorts: the Alzheimer’s Disease Neuroimaging Initiative (ADNI), the Australian Imaging, Biomarker & Lifestyle Flagship Study of Ageing (AIBL), the AddNeuroMed, the Group of Neuropsychological Studies from the Canary Islands (GENIC, from *Grupo de Estudios Neuropsicologicos de las Islas Canarias*), the Japanese ADNI, and the UK Biobank (Figure 1). The description of each cohort is in the Supplementary Material, Section A. A cognitively healthy status was defined based on the absence of dementia, mild cognitive impairment, and other neurological and psychiatric disorders. Further, individuals had to have a clinical dementia rating (CDR) score equal to zero, or mini-mental state examination (MMSE) score ≥ 24, or self-reported good health (this last when available), or ICD-9 or 10 (details on the used ICD codes can is in Supplementary Material, Section B), depending on the available data in each cohort.

**Figure 1.**
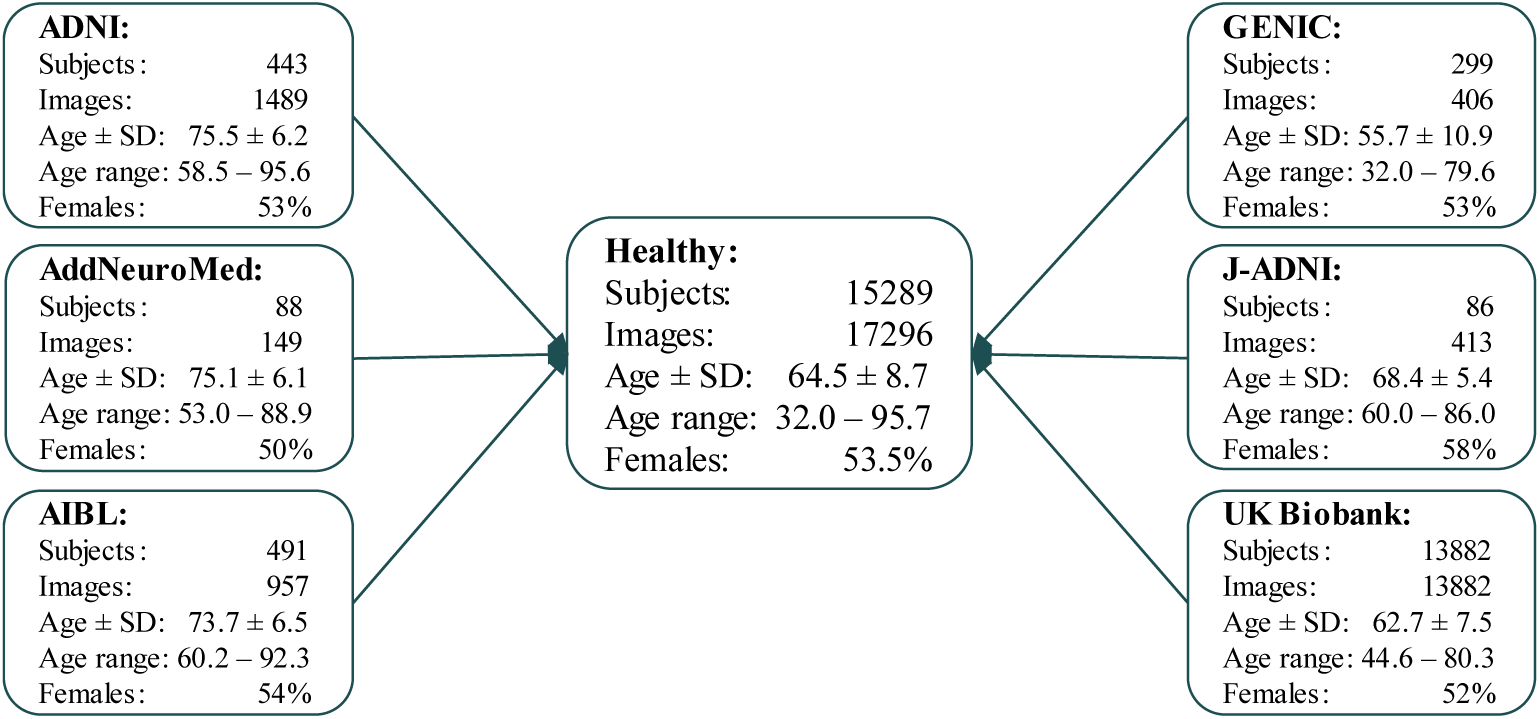
Flowchart describing the data included in this study, which were available in our database system at the time of the study, for the different cohorts. Where age ± SD is the mean age and standard deviation, age ± SD and age range are in years.

### 2.2 CONVOLUTIONAL NEURAL NETWORKS

The implemented CNN uses the deep learning framework PyTorch (Paszke et al., 2019). The network architecture of our model was based on the ResNet architecture (He et al., 2016), with 26 layers in total but with 3D kernels. Each convolutional operation is followed by batch normalisation and a Rectified Linear Unit (ReLU) activation. The network was trained for 20 epochs with stochastic gradient descent and an initial learning rate of λ=0.002 that decreases with a factor of 10 every five epochs. We used five independent models during the CNN development and the trained networks as an ensemble model. Data augmentation included random scaling, cropping offsets, rotations (-5 to 5 degrees), affine, and gamma transformations (ranging from 0.5 to 2). Each image was cropped to a dimension of 80 x 96 x 80 voxels, with 2 x 2 x 2 mm³ resolution, thresholded to the 5th and 95th percentile of the voxel values and scaled so that all voxels’ values were in the interval [-1,1].

We streamline the image pre-processing to improve the model’s accessibility and processing speed. The sole pre-processing step involved a rigid registration (with six degrees of freedom) to the MNI template space using FSL FLIRT 6.0 (FMRIB’s Linear Image Registration Tool). A rigid registration is more straightforward and quicker than an elastic registration. Omitting this step resulted in worse performance on the development set, despite the use of heavy data augmentation (data not shown). Figure 2 shows a schematic representation of the CNN model.

**Figure 2.**
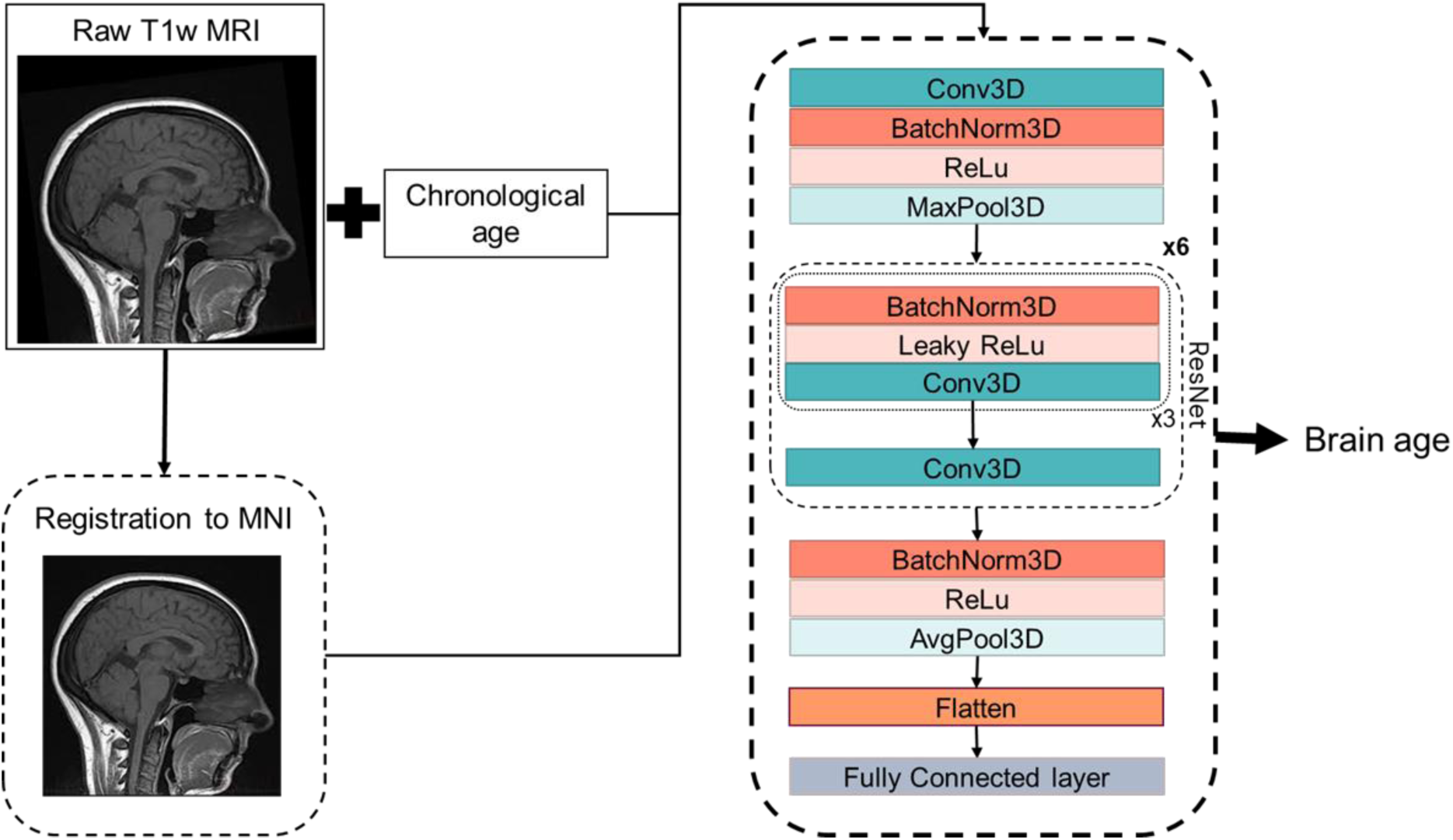
Simplified scheme of our 3D CNN. Where the input is a T1w brain MRI registered to the MNI space, the information on the subject’s chronological age, and the output is the predicted brain age of the subject. Conv3D = 3D convolution: BatchNorm3d: 3D batch normalisation; (Leaky)ReLu: (leaky) Rectified Linear Unit; MaxPool3D: 3D max pooling; AvgPool3D: 3D average pooling; FC: fully connected layer; ResNet: residual network block.

Four separate models were developed in this project: one model based on a hold-out approach of CNN (CNN1) and three models based on a cross-validation approach (CNN2-4). Figure 3 displays the evaluation process’s scheme of our CNN model. Each one of the turquoise rectangles represents 1/10 parts of the primary dataset, composed of 16734 raw MRIs from ADNI, AIBL, GENIC and UKB cohorts. Light blue rectangles indicate the 149 MRIs from AddNeuroMed cohort, whereas lilac rectangles indicate the 413 MRIs from J-ADNI. The CNN1 model is based on a hold-out approach with the training (80%), development (10%) and test (10%) datasets indicated by the arrows. CNN2 and CNN4 models incorporated all data from ADNI, AIBL, GENIC and UKB cohorts in their cross-validation loop, while AddNeuromed and J-ADNI were used for external validation. The CNN3 model was similar to CNN2 and 4, except that AddNeuroMed and J-ADNI were also incorporated in the cross-validation loop – thus, no external datasets were used for model evaluation.

**Figure 3.**
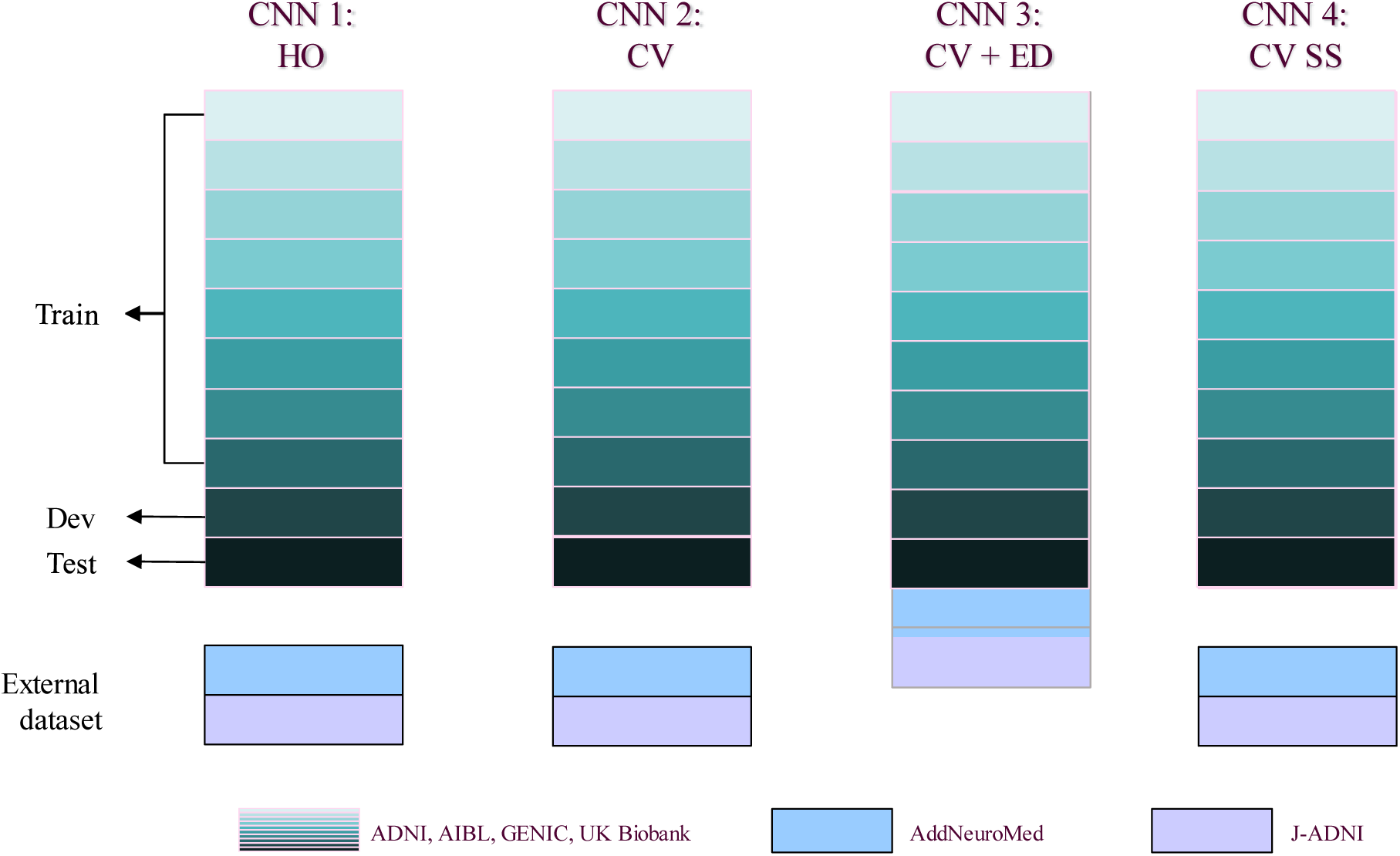
Scheme of evaluation of our CNN model in this study. We primarily used ADNI, AIBL, GENIC, and UK Biobank to develop our model (turquoise scale). CNN1 works in a hold-out approach (data split: 80% train, 10% development, 10% test set, each set of the data indicated by arrows). CNN2 was trained as a 10-folds cross-validation model, using the data of the four primary cohorts (turquoise scale) in the training loop. To evaluate the performance of these two models, we used AddNeuroMed (light blue) and J-ADNI (lilac) as external test sets. In CNN3, we added the external test sets in the 10-fold cross-validation. For comparison reasons, we evaluated our CNN2 scheme with skull stripped T1w MRIs (CNN4). *HO = hold-out; CV = cross-validation; ED = external test set; SS = skull-stripped images*.

#### 2.2.1 Hold-out approach

Our CNN-based model was first trained in a hold-out fashion (CNN1). To not violate the test set and ensure comparability between the models, the primary dataset, composed of 16734 MRIs from ADNI, AIBL, GENIC and UKB cohorts, was randomly split into a training (N*_img_* = 15052, N*_subj_* = 13612, subsequently split into internal validation set for the development of each model) and a hold-out test (N*_img_* = 1682, N*_subj_* = 1503) set. If subjects had undergone multiple scans, all their images were assigned to the same set. The test set was evaluated after a satisfactory performance on the internal validation set to reduce the risk of model overfitting. The data distribution in train, development, and test sets for each cohort can be found in Supplementary Material, Section C. After training the CNN1 model, we applied this model in the AddNeuroMed and J-ADNI cohorts, to assess the model performance and generalizability in external datasets.

#### 2.2.2 Cross-validated approach

To allow comparability to the hold-out model (CNN1), we used the same 16734 MRIs from ADNI, AIBL, Genic, and UK Biobank in the cross-validation approach training loop (CNN2). Stratification by cohort was applied in splitting the 10-fold for training and testing. After a 10-fold cross-validation, the trained model was evaluated in AddNeuroMed and J-ADNI (external cohorts). Furthermore, to ensure that our model’s prediction was based on the brain and not on other features (e.g., head shape), we trained a 10-fold cross-validation model using skull-stripped brain images (CNN4). For CNN4 model images input, Freesurfer 6.0.0 was used to perform skull-stripping, applying the algorithm recon-all and selecting the image generated before brain parcellation (brain.finalsurfs.mgz). Images were motion and bias-corrected, transformed to Tailarach space, intensity normalised, and skull stripped. To reduce the size of the final processed image and for comparative reasons, all skull-stripped images were rigidly registered to the MNI space. Similarly to CNN2, also the skull-stripped CNN4 model was externally validated in AddNeuroMed and J-ADNI. Finally, to ensure the cross-validated model’s usability when including more diverse data, we trained a last model, CNN3, that included all cohorts (ADNI, AIBL, Genic, and UK Biobank plus AddNeuroMed and J-ADNI) to the ensemble of images within the 10-fold cross-validation.

### 2.3 ANALYSES

#### 2.3.1 Model performance

Model performance was assessed using the MAE, defined as:

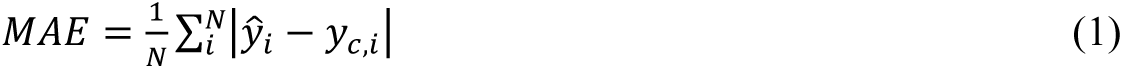

Where *y*_*c,i*_ is the chronological age of participant *i* and *y^*_*i*_ the predicted age. MAE’s values close to zero, indicates model’s good performance, with predicted brain age being similar or almost equal to chronological age but considering aging heterogeneity, is unknown the lower limits for a model’s MAE. Adjusting for age-dependent predicted brain age differences from the chronological age (brain age difference/gap - BAG) is problematic and can artificially inflate model performance (Butler et al., 2021). This can be illustrated with a model that, regardless of input data, only outputs a single predicted age of, e.g., 70. This will yield a suboptimal MAE, but when “correcting” for age, the MAE will be 0 between predicted and chronological age.

#### 2.3.2 Relevant regions for brain age prediction

To explain the model’s brain age prediction, we generated 3D gradient-based saliency maps of each subject using the SmoothGrad (Smilkov et al., 2017) algorithm. Salience maps visualizes the important voxels in individual predictions based on the computation of the gradient of the prediction with respect to the smoothed image. For gradient computation, we used the image with 15% noise added. The 3D gradient maps were averaged through the whole image sample, and only 1% of the higher salient values are shown to verify the most critical regions (Levakov et al., 2020; Mouches et al., 2022). For individual extrapolation, we plotted the 1% normalised higher values of the salience maps and overlayed them onto an arbitrary T1w brain MRI. The salience maps are presented according to their brain age difference in the CNN1 model, calculated from the chronological age, from -8 to +8 yrs. of difference from chronological age.

#### 2.3.3 Differences in cortical thickness across age groups based on chronological and predicted brain age

To analyse the influence of correcting an individual’s brain age in neuroimaging studies, we visualise how brain age predictions are related to cortical thickness values. We ran surface group analysis with QDEC (Query, Design, Estimate, Contrast) in FreeSurfer 6.0.0. We used a smoothing kernel of full width at half maximum of 10 mm, used sex as a covariate and adjusted for false discovery rate at a threshold of 0.05. We grouped subjects of ages 60, 65, 70, 75, and 80 (±1 years) based on chronological and predicted ages. These groups were contrasted to a reference group of 55 ± 1 years-old (based on corresponding chronological or predicted brain age) individuals in the general linear model. Since these groups are of different sizes—and *p*-values are influenced by group size—we present figures overlayed with z-scores.

## 3 RESULTS

### 3.1 MODEL PERFORMANCE

Scatterplots of the brain age predictions on the CNN1 (hold-out) approach for the training and testing set of healthy individuals are shown in Figure 4. The results show that there is a strong correlation between chronological and predicted brain age in training and test sets.

**Figure 4.**
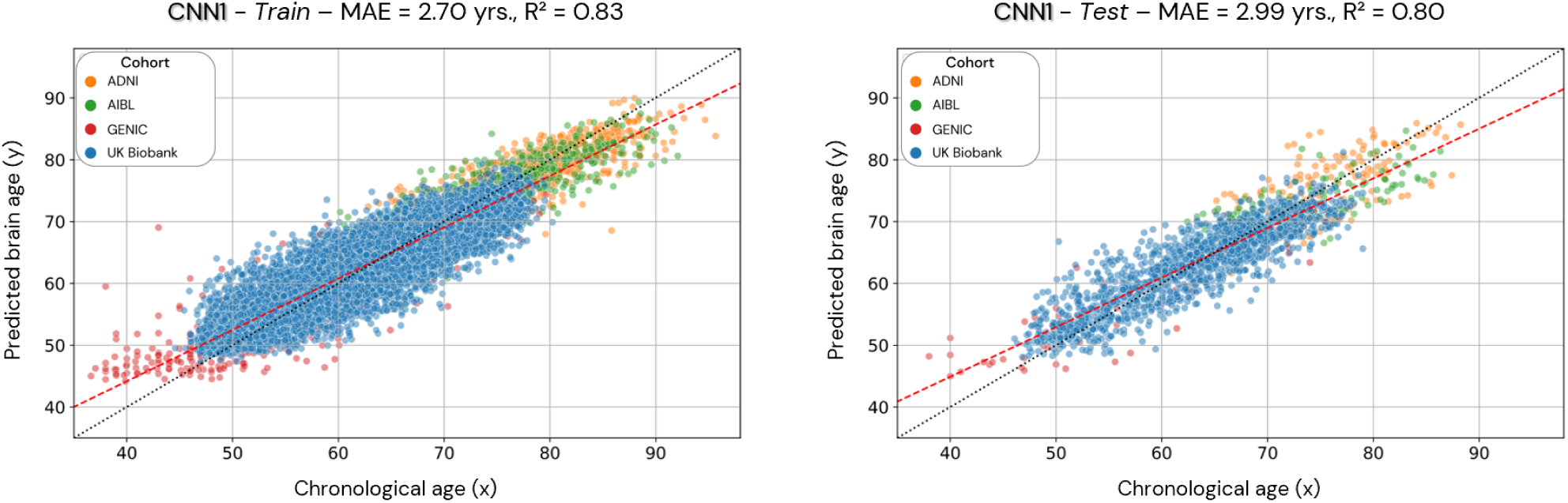
Scatterplots of the predicted brain age in the CNN1 model (hold-out approach) in the training and test datasets. Each coloured dot represents an individual, and each colour is a different cohort. The red line is a linear regression based on the predicted brain age.

Scatterplots of the brain age predictions on the CV approach for the CNN2 (CNN1 cohorts), CNN3 (CNN1 cohorts + J-ADNI and AddNeuroMed), and CNN4 (skull-stripped images from CNN1 cohorts) are shown in Figure 5. For CNN2 and CNN3 models, results show a strong correlation between chronological and predicted brain age with a MAE within the lower range of previously published models. However, CNN4 present a less strong correlation and higher MAE within the presented models in this study.

**Figure 5.**
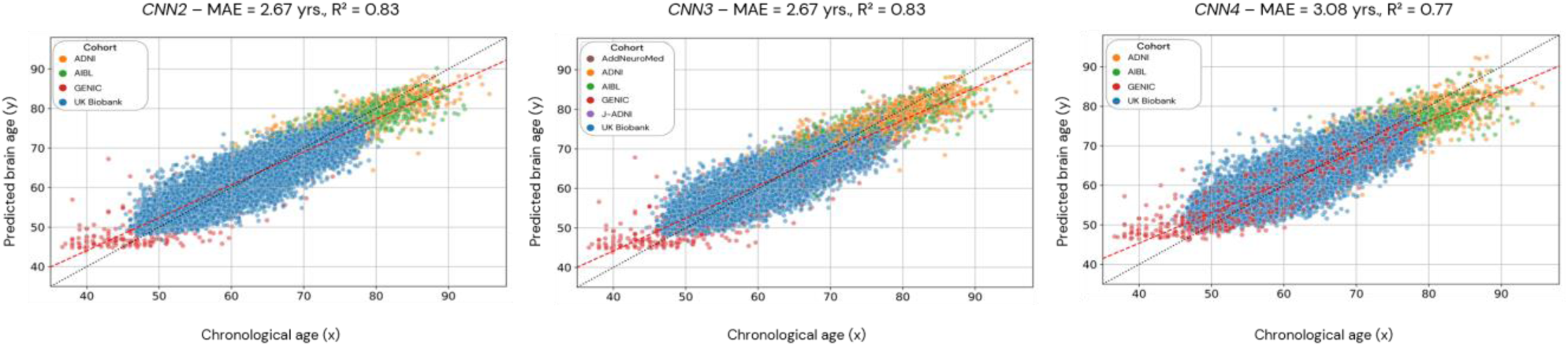
Scatterplots of the predicted brain age in the CNN2, 3 and 4 (cross-validation approach). Each dot represents an individual, and the colour code used for each cohort is presented in the legend. CNN2 was run using 4 cohorts, CNN3 with 6 cohorts, and CNN4 with 4 cohorts but with skull-stripped images. The red line is the linear regression based on the predicted brain age.

Figure 6 (A) illustrates CNN1 (on the left) and CNN2 (on the right) correlations between the predicted brain age (y-axis) by each model in the data from the test set of the CNN1 model, with the chronological age (x-axis). Figure 6 (B) illustrates the correlation between the predicted brain age by CNN1 (x-axis) and CNN2 (y-axis) using the training (on the left) and test (on the right) dataset splits of CNN1. Figure 6 (C) shows the correlations between the brain age difference (BAG) estimated by the two models (CNN1 on the x-axis and CNN2 on the y-axis) in the training and test datasets splits of CNN1.

**Figure 6.**
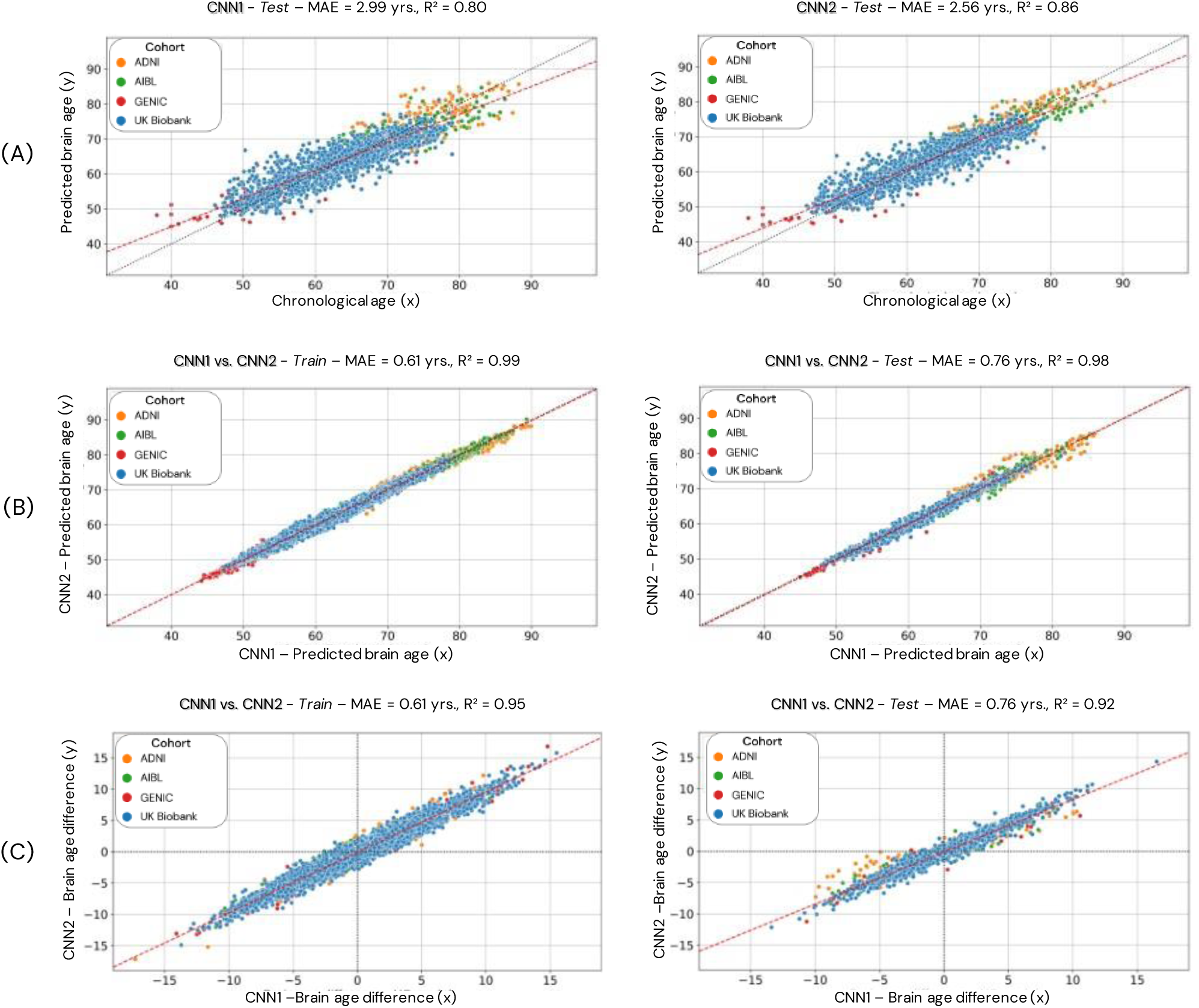
Comparison of the CNN1 (hold-out approach) and CNN2 (cross-validation approach) models. Data used for the comparison is based only on the individuals used in the test dataset of CNN1. In A, we present the test dataset’s predicted brain age (y-axis) for both models correlated to chronological age (x-axis). In B, we present the correlation between the predicted brain age estimated by the two models (CNN1 on the x-axis and CNN2 on the y-axis) in the training (left) and test (right) datasets splits of CNN1. In C, we present the correlation between the brain age difference estimated by the two models (CNN1 on the x-axis and CNN2 on the y-axis) in the training and test datasets.

We also evaluate the performance of our model in the external datasets AddNeuroMed and J-ADNI (CNN1, 2 and 4). The age prediction distribution is shown in Figure 7, and shows the variability in age prediction for each one of the trained models when applied to unseen data (external dataset)Figure 7.

**Figure 7.**
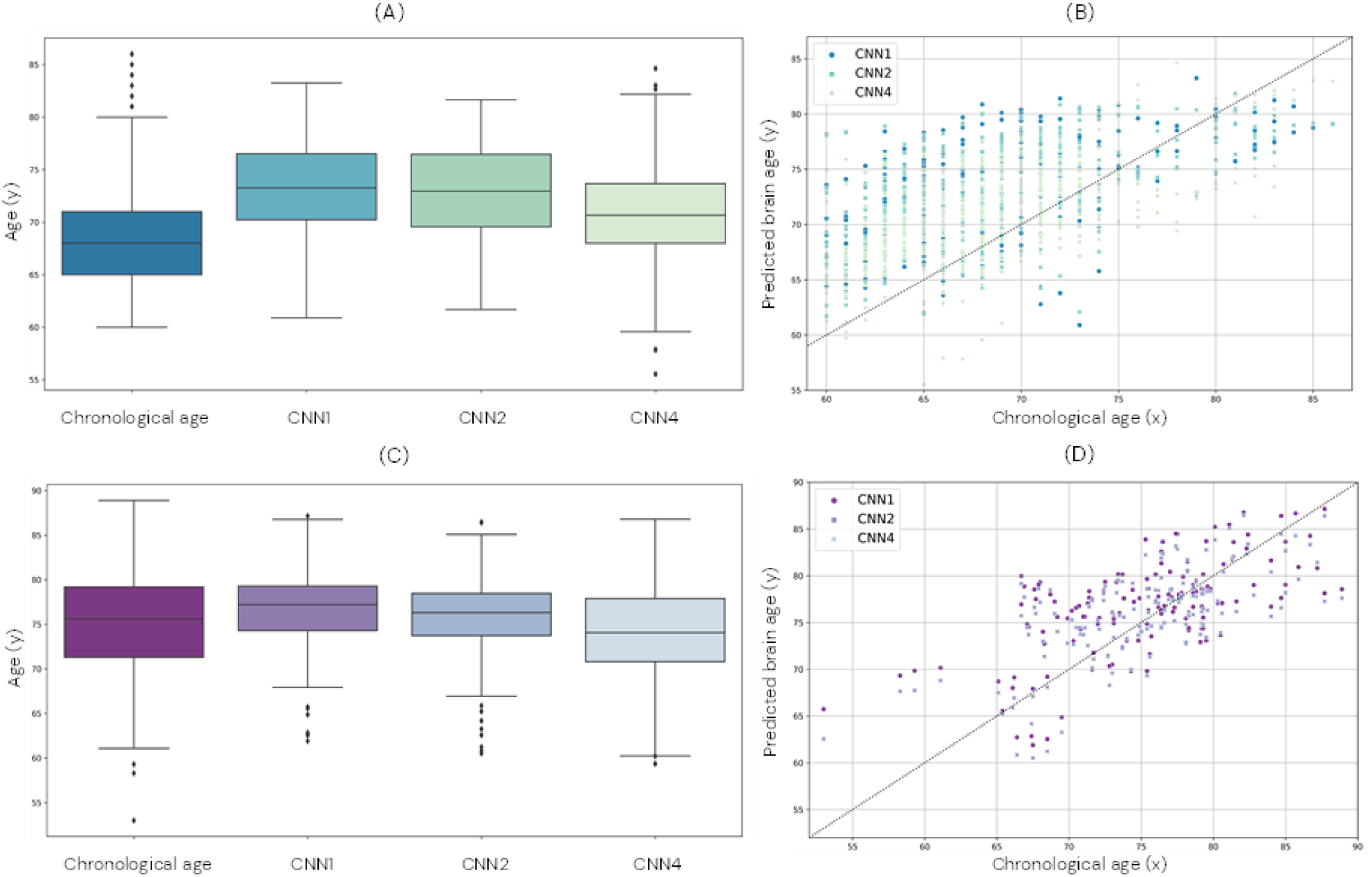
Comparison of brain age prediction distribution for chronological and predicted brain age in the CNN1, 2 and 4 for the AddNeuroMed (A and B) and J-ADNI (C and D) cohort. In A and C, we present the boxplot of the predicted brain age for each model compared to the chronological age. In B and D, the predicted brain age distribution is compared to the chronological age.

The calculated MAE of each cohort for all models is presented in Table 1. Normalized MAE and coefficient of determination for each cohort of the trained models can be found in Supplementary Material, Section D, Table D-1 and D-2.

**Table 1.** Calculated MAE for each cohort for each of the trained models.

| Model | MAE per cohort (years) |  |  |  |  |  |
| --- | --- | --- | --- | --- | --- | --- |
|  | ADNI | AIBL | GENIC | UK Biobank | AddNeuroMed | J-ADNI |
| CNN1<sup>1</sup> | 2.56 | 2.85 | 4.20 | 2.75 | *4.03* | *5.63* |
| CNN2<sup>2</sup> | 2.40 | 2.69 | 3.96 | 2.66 | *3.67* | *5.30* |
| CNN3<sup>2</sup> | 2.41 | 2.69 | 3.98 | 2.67 | 3.02 | 2.25 |
| CNN4<sup>1</sup> | 2.98 | 3.33 | 4.42 | 3.03 | *3.64* | *4.14* |

The numbers in italic (AddNeuroMed and J-ADNI cohorts, for models CNN1, 2 and 4) were calculated as external datasets to the model. <sup>1</sup>Statistical differences were founded (p-value < 0.05) comparing the CNNs absolute error; <sup>2</sup>No statistical differences were founded in the comparison between absolute error of CNN2 and CNN3 (p-value = 0.37). Statistical differences between the models were calculated using Friedmans test with Conover's *post hoc* test, adjusting for multiple comparisons with the Bonferroni method.

To understand how our model performed compared to existing models, we further assessed our brain age models’ performance only within the UK biobank. Then we compared the achieved MAEs with previous studies that evaluated their models in the UK Biobank cohort (Table 2). Our CNN models achieved MAEs ranging between 2.66 and 3.03 yrs. These are very similar to the MAEs achieved by CNN models developed in previous studies (ranging between 2.13 and 4.36). We also present the coefficient of determination between predicted brain age and the identity line for all the available studies.

**Table 2.** Comparison of our models MAE with the literature.

| Study | Model | Modality | Pre-processing | MAE | R<sup>2</sup> |
| --- | --- | --- | --- | --- | --- |
| Dartora et al. | CNN1 | T1 | Rigid reg. | 2.75 | 0.88 |
| Dartora et al. | CNN2 | T1 | Rigid reg. | 2.66 | 0.89 |
| Dartora et al. | CNN3 | T1 | Rigid reg. | 2.67 | 0.89 |
| Dartora et al. | CNN4 | T1 | Bias-field and motion cor., Skull-strip., Rigid MNI reg. | 3.03 | 0.85 |
| Bintsi et al. (2020) | CNN (3D ResNet) | T1 | Skull-strip., non-linear MNI reg. | 2.64 | 0.77 |
| Bintsi et al. (2020) | CNN (patches) | T1 | Skull-strip., non-linear MNI reg. | 2.13 | 0.85 |
| Kolbeinsson et al. (2019) | CNN | T1 | Skull-strip., non-linear MNI reg. | 2.58 | - |
| P. K. Lam et al. (2020) | R-CNN | T1 | Bias-field cor., Skull-strip., Rigid MNI reg. | 2.86 | 0.87 |
| P. Lam et al. (2020) | CNN | T1 | Bias-field cor., Skull-strip., Rigid MNI reg. | 4.36 | 0.66 |
| Gupta et al. (2021) | CNN (slices) | T1 | Bias-field cor., Skull-strip., Rigid MNI reg. | 2.82 | - |
| Dinsdale et al. (2021) | CNN (Female population) | T1 | UK Biobank pipeline w/ linear registration to MNI | 2.86 | 0.87 |
| Dinsdale et al. (2021) | CNN (Male population) | T1 | UK Biobank pipeline w/ linear registration to MNI | 3.09 | 0.86 |
| Jonsson et al. (2019) | CNN (transfer learning) | T1 | Bias-field cor., Skull-strip., Dartel MNI reg., tissue maps | 3.63 | 0.61 |
| Peng et al. (2021) | SFCN | T1 | Bias-field cor., Skull-strip., Rigid MNI reg. | 2.14 | 0.39 |
| Peng et al. (2021) | CNN | T1 | Bias-field cor., Skull-strip., Rigid MNI reg. | 2.38 | - |

CNN: convolutional neural network; CNN1: our CNN model in a hold-out approach; CNN2: our CNN model in a cross-validation approach; CNN3: our CNN model in a cross-validation approach and including two cohorts in the dataset; CNN4: our CNN model in a cross-validation approach, using skull-stripped images; PCA: principal component analysis; R-CNN: recurrent CNN; SFCN: simple fully convolutional network; R<sup>2</sup>=coefficient of determination. Dartora et. al. refers to the current work.

To understand the noise levels from our models and their ability to capture subtle changes as a result of the ageing process, we plotted longitudinal trajectories for the participants with more than one-time point (Figure 8).

**Figure 8.**
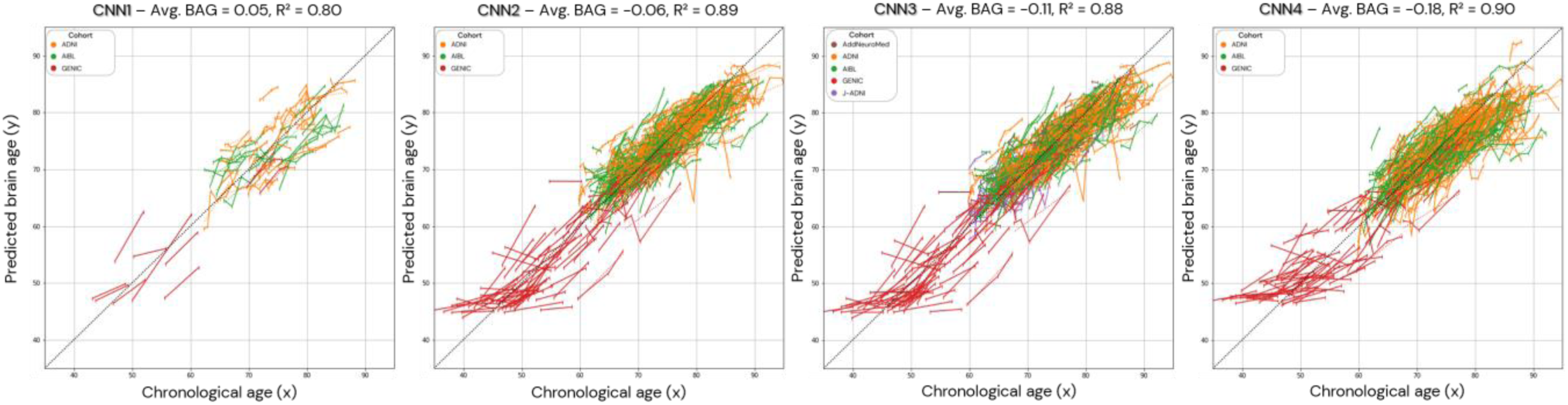
Brain age prediction in longitudinal trajectories for all the models. The average brain age gap/difference (Avg. BAG) in the presented population was calculated and shows a trend towards zero to the brain age difference between predicted and chronological age. In CNN1, only the individuals with longitudinal data in the test set are presented.

### 3.2 RELEVANT REGIONS FOR BRAIN AGE PREDICTION

For the explicability of our model, the salience map of each individual prediction was generated. The averaged overlayed salience maps for each CNN model are presented in Figure 9. Complementary view of salience maps slices is presented in Supplementary Material Section E, Figure E-1.

**Figure 9.**
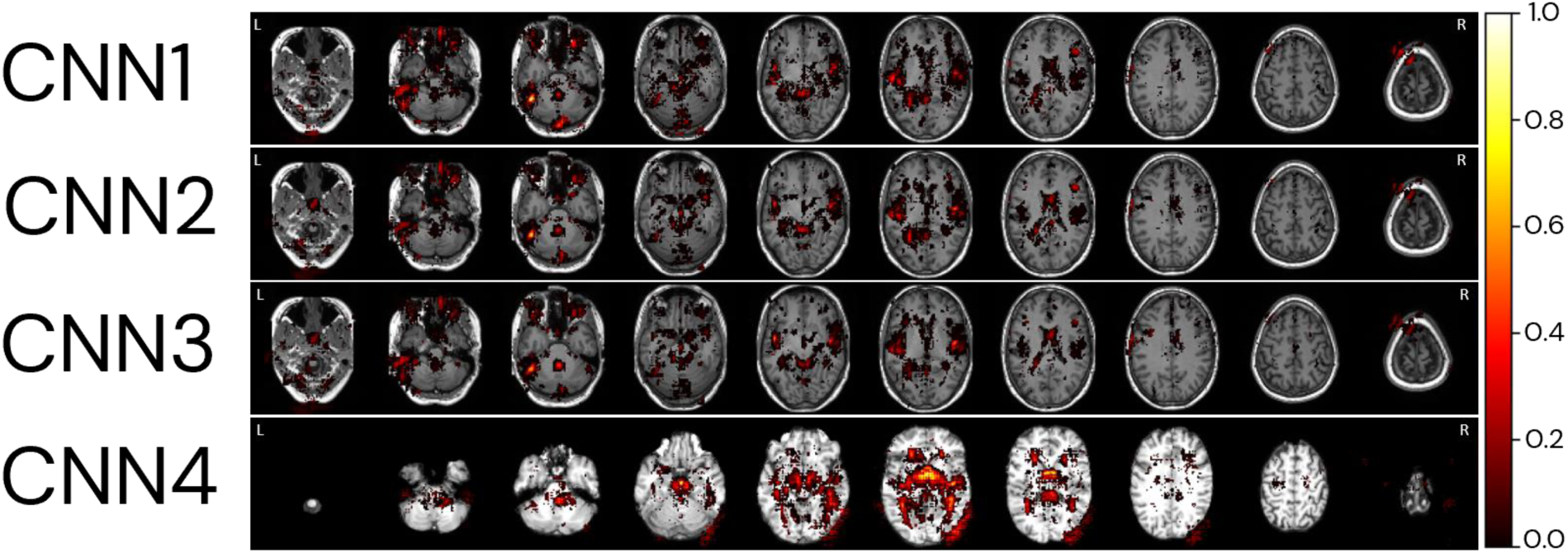
Relevant regions for age prediction. The absolute values of the salience maps for each model were averaged through the whole MRI sample and normalised between 0 and 1 for better visualisation. The SmoothGrad method was used to generate all salience maps. Overlayed salience map colours are normalised for each individual between 0 and 1.

To identify regions of importance for predicting the biological brain age, 18 individuals were randomly selected according to the following criteria: being in the test dataset of the CNN1, being between 64 and 66 yrs., having an age range difference between chronological and predicted brain age in the CNN1 model between -8 and +8 yrs. (Figure 10). The brain age gap for each individual, predicted in each model, is presented in Supplementary Material, Section F, Table F-1.

**Figure 10.**
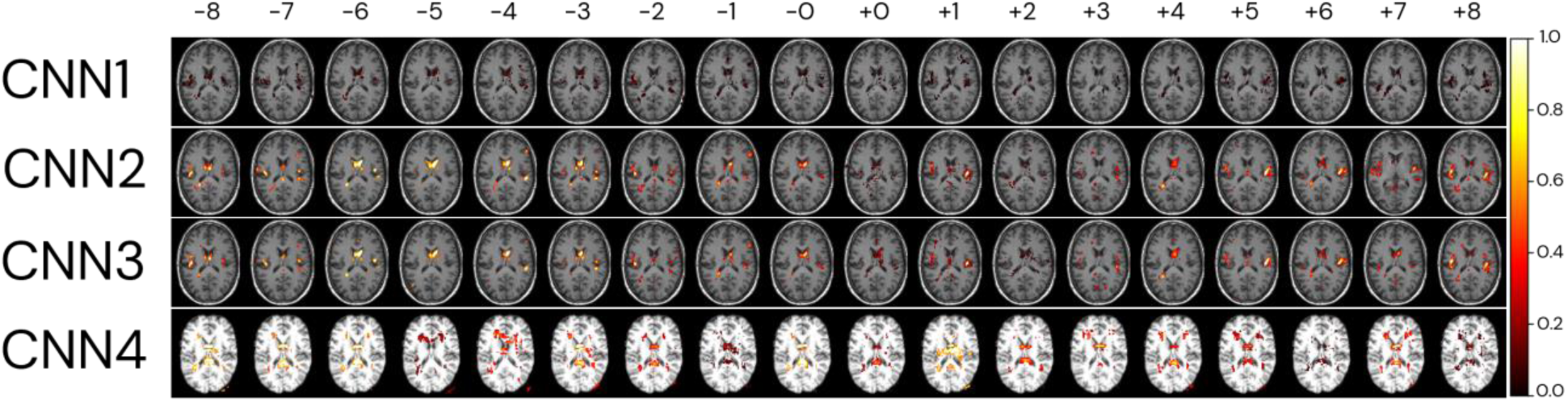
Relevant regions for the brain age prediction based on 1% of the salience maps of individuals with an average age of 65 yrs. overlayed to a random T1w MRI sample for each of the four CNN models. The individuals were randomly selected in the test dataset used in the CNN1, according to their brain age difference and proximity to the average age of our sample (65 yrs.). Each row represents the salience maps of one CNN model, where the columns show the same individual’s salience map in each of the four models. The SmoothGrad method was used to generate all salience maps. The brain age difference is shown in the top row of the image, going from -8 to +8 yrs. of difference. Overlaid salience map colours are normalised for each individual between 0 and 1.

### 3.3 DIFFERENCES IN CORTICAL THICKNESS ACROSS AGE GROUPS BASED ON CHRONOLOGICAL AND PREDICTED BRAIN AGE

Surface group analysis for cortical thickness was run in all available images with QDEC in Freesurfer 6.0.0. Individuals with 60, 65, 70, 75, and 80 (±1 years) chronological and biological (predicted) age were grouped and compared with a group of 55±1 years-old individuals. The analysis used sex as a covariate and was adjusted for a false discovery rate with a threshold of 0.05. Figure 11 shows the age-related differences in cortical thickness for chronological and biological (predicted) age groups by the CNN2 model. The number of individuals in each age group is presented in Supplementary Material, section G, Table G-1.

**Figure 11.**
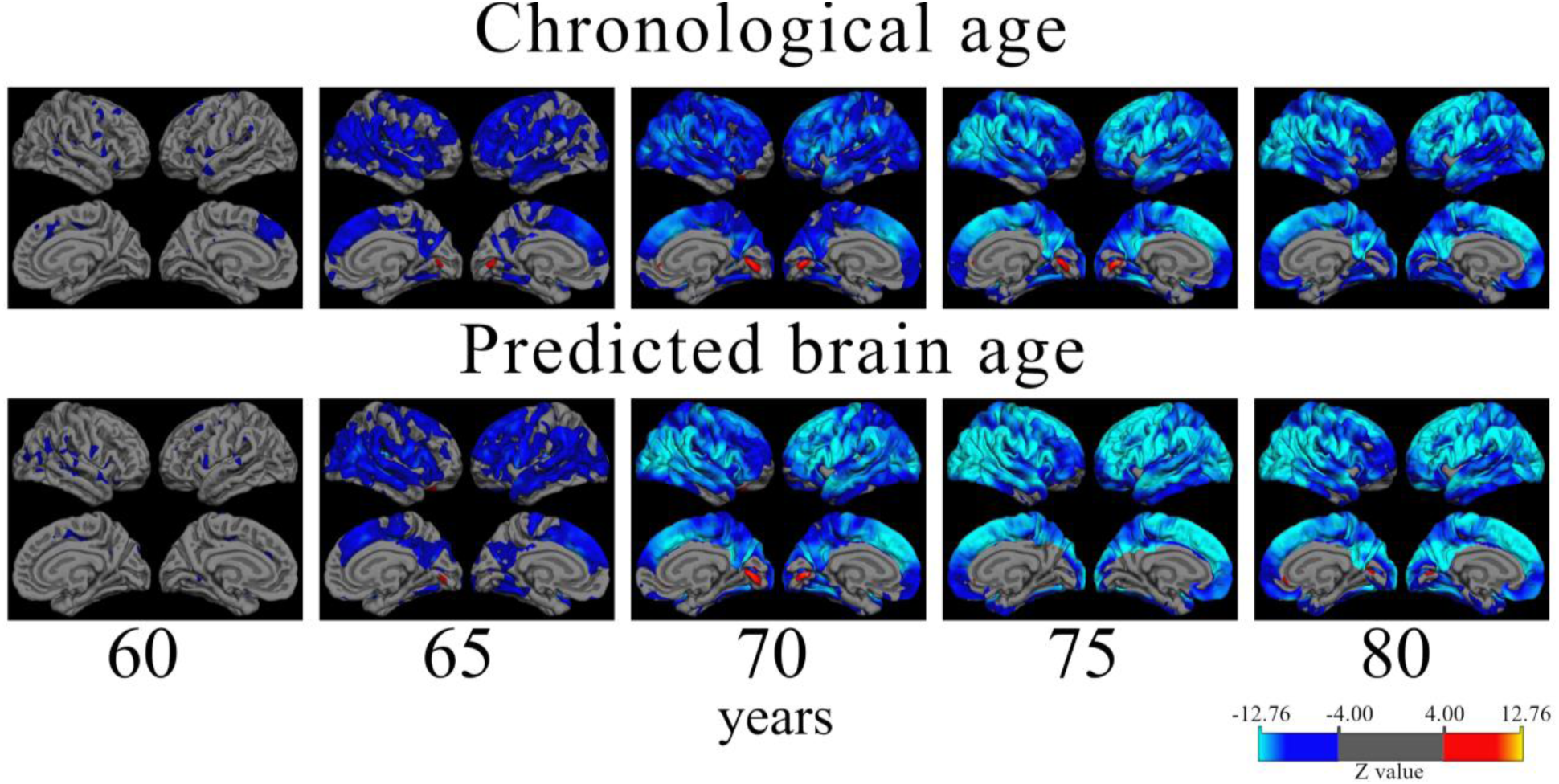
Atrophy patterns in aging according to chronological and predicted brain age in the cross-validation approach (CNN2). Analysis shows cortical thickness z-stat maps corrected for false discovery rate with a threshold of 0.05 of cross-sectional differences between a reference group of 55 (±1.0) years old and older subjects. Individuals were grouped by age ±1.0 years. The QDEC analyses were done with a 10 mm smoothing kernel and sex added as a covariate. Blue colours represent lower cortical thickness (increased atrophy) in the age group compared to the 55-year-old group, and red colours represent an increased cortical thickness compared to the younger group. It is possible to verify that the atrophy patterns start in parietal-frontal regions from the 65-years old group. The approach also captures the increased thickness (in red) around the lingual gyrus.

## 4 DISCUSSION

In this study, we developed a CNN-based model using only one pre-processing step (rigid registration to MNI space) to predict the person’s biological brain age from T1w images, to be easily implemented and used. Further, the study was performed as an effort to attend to the highlighted points related to an “ideal” image database (which includes diverse image data) to develop brain age prediction models. Finally, our model evaluation was performed using different approaches – hold-out (CNN1) and cross-validation (CNN2) – and generalizability was tested in external datasets. The usability (CNN3) of the model was also assessed by adding more data (from two external datasets) in the training loop, and to ensure that the previously trained models were using the brain signal for age prediction, we also trained the model using skull-stripped images (CNN4).

A key strength of this work is the use of minimally processed images as input for the CNN model, which makes it feasible for implementation in research and, in the long term, clinical settings. Our model requires only the registration to MNI space, which typically takes a few seconds and can be easily performed using an open brain image processing software such as FSL or FreeSurfer. Additional pre-processing steps would increase the likelihood of image exclusion during quality control and pre-processing. This would limit the model’s performance as well as the possibility of using cohorts with a small sample size. CNN models can learn from the image data, including structure shape, which may not be captured by summary metrics such as volume or segmented tissue maps, without requiring pre-segmented data (Liang et al., 2019; Niu et al., 2020). Our focus on a one-step pre-processing, using a rigid registration to the MNI space template previously implemented by Cole et al. (2017), is to ensure the accessibility to our model in future research.

This study includes cohorts to attend to the highlighted points related to, what we believe to be, an ideal image database to develop brain age prediction models. We used detailed information and clinical data for inclusion criteria of many individuals (more than 16000 T1w MRIs) from different parts of the world (Asia, Australia, Europe, and America) and with a diverse number of MRI scanners covering 1.5T and 3T scanners, which increases the model usability due to its generalisation to new unseen data (Mårtensson et al., 2020). We also used all the longitudinal data available and showed that our model presents a reasonable age prediction congruent with the individual timeline**Error! Reference source not f ound.**. Also, our focus on the cognitive “healthy” status, rather than on the overall health status (for example, excluding from the training set age-related diseases that affect the body’s organs other than the brain) is a strength of our study as it makes a clear separation between the outcome (brain age) and what leads to that outcome (risk factors, e.g., chronic cardiometabolic disease and risk factors, affective/mood disorders, etc). Further, to validate the generalizability of the models, we used external datasets of cognitively unimpaired individuals from two cohorts: AddNeuroMed, a cross-European study designed to find biomarkers for Alzheimer’s disease, and J-ADNI, the Japanese version of the ADNI dataset. We chose to use these cohorts because of differences in age distribution, e.g., AddNeuroMed average age is around ten years older than the average of our total sample, different ethnicity, e.g., our sample is composed mainly of European and North-American individuals, and both cohorts mostly use images acquired in 1.5T scanners, whereas the training dataset was based mainly on 3T MRI, i.e., 96.2 % of all images used for training and testing in the CNN1 approach.

Due to the complexity and time-consuming nature of the training networks with large amounts of data, the hold-out method used in CNN1 is the most common approach in the literature (Cole et al., 2017; Dinsdale et al., 2021b; Gupta et al., 2021; Jonsson et al., 2019; Kolbeinsson et al., 2021; Lam et al., 2020; Peng et al., 2021; Ren et al., 2022). Our CNN1 model achieved an MAE of 2.99 yrs. in the test set, which agrees with the MAE reported in the literature for hold-out test sets, using 1 to 3 cohorts (MAE of 2.14 – 4.65 yrs.) (Cole et al., 2017; Dinsdale et al., 2021b; Gupta et al., 2021; Jonsson et al., 2019; Kolbeinsson et al., 2021; Lam et al., 2020; Peng et al., 2021; Ren et al., 2022). Our calculated MAE is also in the range of the available MAEs of the CNN models (Bintsi et al., 2020; Dinsdale et al., 2021b; Gupta et al., 2021; Jonsson et al., 2019; Kolbeinsson et al., 2021; Lam et al., 2020; Peng et al., 2021) available in the literature that used hold-out approaches and only data from the UK Biobank but performing several pre-processing steps as opposed to our CNN model. The current debate surrounding the development of brain age models lacks a thorough evaluation of CNN-based brain age models across different external datasets. In such scenario, it is difficult and relatively unfair to compare the performance of different models not using the same evaluation dataset (Sajedi and Pardakhti, 2019). To overcome such a challenge, we compared the performance of our CNN in its different approaches related to the two used external datasets, AddNeuroMed and J-ADNI, showing that CNN1’s MAE performance is still within a range of 1.5 years when compared to the other CNNs.

Deep learning models, like CNNs, tested in out-of-distribution data, results in performance dropping, increasing underestimation in new unseen data, e.g., clinical data. However, including more diverse and variable data to the model’s training, increases models’ robustness/reliability (Mårtensson et al., 2020). One way of including variability in the model is running it in a cross-validation fashion. We have done this in CNN2, where the same data used for training, validating, and testing CNN1 was used for training a 10-fold cross-validation model (CNN2). In general, the CNN2 model had a better performance than CNN1. The correlation with chronological age was higher, the MAE was smaller in CNN2 in all cases of age prediction (training and test sets), and calculated brain age difference (BAG) when comparing both models. Comparing the performance in the different cohorts used in the development of this work, we see a tendency of individual smaller MAEs in CNN2. To confirm the robustness of the model, CNN2 was also evaluated in external datasets, confirming the decrease in underestimation, proved by the increase of the model’s performance in AddNeuroMed and J-ADNI cohorts when compared to CNN1. Even though CNN1 and CNN2 have similar performance in age prediction, the increased variability in the model’s training reduced the average and calculated MAE for each cohort. This is further proved by CNN3, where CNN2 and CNN3 performed similarly (p>0.05). Both models presented the same MAE of 2.67 years and a coefficient of determination of 0.83 but different performances in out-of-distribution data. Specifically, evaluating the predictions in the J-ADNI cohort in CNN3, it is possible to verify that adding the cohort to the training set decreased the predicted MAE for this cohort, going from an MAE of 5.63 in CNN1 to 2.25 in CNN3. This reinforces our hypothesis that the small number of data does not increase the absolute error across all the cohorts (AddNeuroMed and J-ADNI represent ∼3% of the total amount of images used in CNN3) but increases the performance in a specific cohort. This increases the model’s usability, as data with more variability will be included in the training set, ameliorating the age prediction in external cohorts (Mårtensson et al., 2020). Therefore, additional tests with a greater variety of cohorts are necessary to understand how wide the model’s generalizability is and how it can change by adding more data to the training set.

To ensure that our model was using only brain signals from the T1w MRI and that predictions were not depending on the head morphology or bone tissue, we trained and evaluated CNN4. This model is comparable to CNN2, apart from that skull-stripped brain images were used as input. Regarding performance, CNN2 presented a smaller MAE and better correlation to the chronological age than CNN4. The same tendency is also present in evaluating data only from the UK Biobank. This shows that our CNN works better with minimally pre-processed brain MRIs when compared to heavily pre-processed images. However, for evaluating the external dataset, J-ADNI, CNN4 presented a smaller MAE than CNN2. We hypothesise that using heavily pre-processed images as in CNN4 could remove or decrease the effects of bias field and skull size/format and partially mitigate the inhomogeneity present in the external dataset.

The lower limit of the MAE score is unknown and depends on both the inter-subject variability and age distribution of both training and test datasets. By using stricter exclusion criteria for what is considered “cognitively healthy”, the variability and theoretical MAE lower bound decrease. This makes comparisons between studies of models evaluated on different datasets challenging. Several studies have trained and evaluated their model on the UK Biobank cohort, which enables rough comparisons. However, this restricts the model to cross-sectional image data (at least for the first wave of the UK Biobank data) from a “homogeneous” population from the United Kingdom acquired in standard equipment (Siemens Magnetom Skyra Syngo MR D13) with 3T MRI following the same protocol, which is not the reality for datasets and clinical/research settings. Also, the performance of CNNs trained on medical images from one cohort may produce systematically different predictions on images outside the training data distribution (Mårtensson et al., 2020). Comparing our results with the literature applied only to UK Biobank images, we observed that using several time-consuming image pre-processing steps, none of the models achieved an MAE smaller than 2.13. The CNN4, which uses skull-stripped images, showed the worst performance within our different approaches using the same CNN architecture. For a more accurate comparison of the model’s performance using the MAE metric, normalised MAE should be used. However, not all the selected papers for comparison presented the average age of the used subgroup of UK Biobank data, limiting the calculation of a normalised MAE. For future comparisons to our work, the normalised MAE for all four different approaches is presented in Supplementary Material, Section F.

Essentially, all our trained models showed MAE levels comparable to those reported in previous literature, which is typically in the range of 2.13 – 6 (Baecker et al., 2021b, 2021a; Bintsi et al., 2020; Cole, 2020; Dinsdale et al., 2021a; Gupta et al., 2021; Jonsson et al., 2019; Kolbeinsson et al., 2019; Lam et al., 2020; Sajedi and Pardakhti, 2019; Tanveer et al., 2022). This indicates that our models have a good performance with the advantage of requiring only one pre-processing step.

A “perfect” model for brain age prediction in cognitive unimpaired individuals should show smooth and non-declining trajectories within the same individual at different time points, assuming that a healthy person’s brain age does not vary rapidly or decreases/increases substantially. Visually, our CNN model in different approaches seems to generate smooth predictions that increase with chronological age, with some noisy predictions deviating from the trajectory. For quantitative analysis, we also calculate the mean age gap (BAG) of all individuals, the mean squared error and R^2^. The models present mean brain age differences between chronological and predicted brain ages smaller than 0.5 yrs. However, the predictions also shows a potential confounder in subsequent analyses of brain age predictions: the noise. As it can be observed, for some subjects, fluctuations between the brain age differed some years between two scanning sessions that were one year apart. It seems unlikely that this is a biological phenomenon but instead attributed to the input data being noisy or low quality. Most datasets do not have abundant longitudinal data to sort “bad” predictions from “good” predictions. If the data and group sizes are extensive, some level of noise is acceptable and might not affect the interpretation of the results. The maximum average difference between chronological and brain age of all longitudinal plots was -0.11, with R^2^’s higher than 0.88. However, this effect can have a non-negligible impact when analysing data sets with small group sizes or running longitudinal analyses with few follow-up scans. This is important to remember when conducting future studies related to brain age, for example, when investigating the association between brain age and neurological disorders with a low prevalence in the population.

The analysis of important brain regions for prediction was realised qualitatively by the plots of the salience maps. In general, the averaged salience map throughout the whole image sample shows to agree with the highlighted regions in the analysis of the brain age difference (BAG) for the same chronological age (65 years). The analysis of brain age differences (BAG) showed similar important regions for prediction, with variations in importance intensity between them. We hypothesise that the salience map shows negligible importance for these specific regions due to the fewer images used to train the CNN1. It is important to highlight that the brain age differences shown in the top row of Figure 10 are based on the CNN1 predictions for the plotted individuals. In general, the CNN4 model shows brain age differences with higher variability than compared with the other three models (CNN1-3). More studies are necessary to understand how this model presents more significant variation, using the same image data but with more pre-processing steps. The intensity of importance (higher importance showed in yellow colours) also shows to be more significant to predict age as younger (e.g., -6 yrs.) than chronological age, mainly close to the ventricles (Bintsi et al., 2021) and insular cortex (Lee et al., 2022). For prediction as older, greater importance is given to the right side of the insular cortex (Lee et al., 2022) as frontal-occipital regions. The salience maps show regions symmetrical and asymmetrical, mainly on the left side of the brain (Roe et al., 2021) and around the ventricles (Bintsi et al., 2021), as important for age prediction. Left brain asymmetries with aging are a typical pattern found in aging studies of cortical thickness (Frangou et al., 2022; Koelkebeck et al., 2014), cortical volume and surface area (Koelkebeck et al., 2014). In agreement with Lee et al. (2022), who plotted salience maps for different decades, regions with a higher contribution for age prediction were in the insular cortex (from 30 – 50 yrs. groups), ventricular boundary (50-60 yrs. group), and cerebellum (90-100 yrs. group).

Interestingly, regions around the eyes were selected in the three non-skull-stripped models (CNN1-3). We hypothesise that changes in the soft tissue and liquid surrounding the orbital space, as the bony orbit, which has differences in sex (i.e., men usually have greater skeleton size than women) (Erkoç et al., 2015), as well as general dimensions in orbital structures (Rana et al., 2022), could be being used in the model to predict age. For the model using skull-stripped brain images as input (CNN4), right regions close to the cerebellum and occipital lobe, outside the brain, were selected as important for prediction. We believe that the increased noise used for the SmoothGrad in a region that could have a higher neurodegeneration load could be leading to prediction importance outside the brain in this model. More studies are necessary to understand the highlight of this region for the prediction. However, we believe this could be an artefact generated by the SmoothGrad method due to the addition of noise in the image for the construction of the maps. Future studies could use different methods to generate salience maps and use different methods to generate skull-stripped images to verify if this outstanding region (right outside the skull region) continues to be highlighted for prediction in this model.

One of the theoretical uses of biological age is for age correction in neuroimaging studies. The hypothesis behind it is that correcting neuroimaging studies for the biological (predicted) age of the individuals will better handle the heterogeneity that we see in aging, incorporating diversity in longitudinal brain trajectories due to lifestyle, environmental or even biological factors (Cole et al., 2019; Tian et al., 2023). For the analysis of chronological and biological (predicted) age atrophy pattern differences, we can observe differences mainly from the older groups (as of 70 yrs.). In the comparison of the group of 70 yrs. in the chronological and biological (predicted) brain age, atrophied areas of higher statistical significance are present in the mid-frontal to parietal-occipital regions of the group based on biological age. This agrees with the work of Thambisetty et al. (2010), where an anterior-posterior gradient in age-related brain atrophy was found, with frontal-parietal regions showing a greater decline. The groups with 75 and 80 yrs. have more similar atrophy patterns, with higher spread to the parietal lobe in the biological (predicted) brain age. Interestingly, a region of greater thickness in the oldest groups, compared to the reference group (55 yrs.), was found in the primary visual cortex, located in the calcarine sulcus. The shrinkage of the visual cortex is still widely discussed in the literature, with a handful of studies showing from cortical thinning of the visual cortex to the sparing of this region. These studies suggest that the visual cortex thickness is use-dependent instead of age-related (Burge et al., 2016; Griffis et al., 2016; Jorge et al., 2020). We hypothesise that this can be a cohort confounding effect, even individuals of 75 yrs. being present in all cohorts, but in larger amounts in ADNI and AIBL. Differences in atrophy patterns between individuals grouped by their chronological and biological (predicted) brain age need to be further studied. However, our results already show different tendencies in atrophy patterns between them. Correcting for biological (predicted) brain age in neuroimaging studies could be one step further in understanding heterogeneity present in aging and be used in early diagnosis of neurological diseases, prognosis and even monitoring of treatment response, being one step further to precision medicine.

Some limitations need to be acknowledged. The large dataset in the current study hindered the possibility of performing extensive quality control. For the CNN model, this would mean inspecting that the performed rigid registration was adequate. The random cases we inspected suggested that the overall quality of the segmentations was sufficient. However, tools to automate the quality control process – such as Brusini et al. (2020) and Klapwijk et al. (2019) – will be necessary for future studies on this data size. Regardless of the lack of extensive quality control of the images, our model showed robust findings with only slightly worse performance when compared to previously published works.

Studies also need to evaluate how the inclusion of different populations influences brain age prediction models based on minimally processed MRIs. However, the training model likely needs to include more diverse data, i.e., different types of cohorts, e.g., from Asia, Africa, and Latin America, as well as a broader range of scanners (from 1.5T to 7T) as well as imaging protocols (Mårtensson et al., 2020). Further, the differences in the distribution of chronological age and the number of subjects in each cohort can lead to overfitting some cohort-specific information and characteristics (e.g., the model could learn that images from GENIC are generally in the lower age span). However, the large training set, heavy data augmentation, and only running 20 epochs in training helped minimise the possibility of overfitting.

Even though we defined the cognitively “healthy” status as consistently as possible across the cohorts, some variation exists but we acknowledge the clinical and cognitive assessments relied on similar procedures across cohorts. Finally, it is worth noting that there might be sociodemographic differences between the cohorts since the recruitment of participants happened in different geographical areas (J-ADNI: Japan, AIBL: Australia, ADNI: North America, and UK Biobank, GENIC, and AddNeuroMed: Europe). However, this is not necessarily a limitation but rather a strength. Indeed, the developed algorithm could be applied in future research, in which the biological age of the brain is a focus, to some extent independent of the cohort characteristics, thus increasing the generalizability of the model. Future studies need to further test this hypothesis and the impact of different cultural backgrounds on the estimation of brain age.

## 5 CONCLUSION

In this study, we developed a CNN-based model to predict biological brain age using raw T1w MRI registered to the MNI space, with the goal of accessibility and simplicity in implementation. The model was systematically evaluated using different approaches, comprising several datasets of cognitively healthy individuals with different scans and population characteristics as well as using cross-sectional and longitudinal data. Our CNN-based model provides accurate results compared to state-of-art methods. The generalizability and usability of the model were tested using external datasets with different demographic characteristics, MRI protocols, and MRI scanners, proving the robustness of the model. In addition, we present the important regions for brain age prediction. We also provide indicators for the use of biological (predicted) brain age as a metric for age correction in neuroimaging studies as an alternative to the traditional chronological age based on the differences in cortical atrophy. Finally, the model’s code and trained CNN weights are made publicly available for the research community to quickly implement and use in their research to study ageing and age-related brain disorders.

## 6 DATA AVAILABILITY

The complete code for using the CNN is available at https://github.com/westman-neuroimaging-group/brainage-prediction-mri. We include all the developed models and their trained weights for researchers to apply on their own neuroimaging data sets (with and without skull-stripped T1w MRI brain images) and scripts for training their model on other datasets. When new image data is available from different cohorts, the CNN will be re-trained, and trained weights and performance of the model will be updated in the page.

## ACKNOWLEDGEMENTS

The authors wish to thank the Swedish Research Council (VR), the Strategic Research Programme in Neuroscience at Karolinska Institutet (StratNeuro), the Center for Innovative Medicine (CIMED), the Foundation for Geriatric Diseases at Karolinska Institutet, the regional agreement on medical training and clinical research (ALF) between Stockholm County Council and Karolinska Institutet, the Swedish Brain Foundation, the Swedish Alzheimer Foundation, the Åke Wiberg Foundation, the Olle Engkvist Byggmästare Foundation, the joint research funds of KTH Royal Institute of Technology and Stockholm County Council (HMT), Swedish Parkinson Foundation, King Gustaf V:s and Queen Victorias Foundation, David and Astrid Hageléns Foundation, Loo and Hans Ostermans Foundation, Vinnova and Digital Futures for financial support, and the NIA-supported Collaboratory on Research Definitions for reserve and resilience in cognitive ageing and dementia. The Titan X Pascal used for this research was donated by the NVIDIA Corporation.

This research has been conducted using the UK Biobank Resource under Application Number 30172. The UK Biobank study was approved by the National Health Service National Research Ethics Service (11/NW/0382). More information about UK Biobank is available at www.ukbiobank.ac.uk.

The AddNeuroMed study was supported by InnoMed, (Innovative Medicines in Europe), an Integrated Project funded by the European Union of the Sixth Framework program priority FP6-2004-LIFESCIHEALTH-5, Life Sciences, Genomics and Biotechnology for Health.

Data collection and sharing for this project was funded by the Alzheimer’s Disease Neuroimaging Initiative (ADNI) (National Institutes of Health Grant U01 AG024904) and DOD ADNI (Department of Defense award number W81XWH-12-2-0012). ADNI is funded by the National Institute on Aging, the National Institute of Biomedical Imaging and Bioengineering, and through generous contributions from the following: AbbVie, Alzheimer’s Association; Alzheimer’s Drug Discovery Foundation; Araclon Biotech; BioClinica, Inc.; Biogen; Bristol-Myers Squibb Company; CereSpir, Inc.; Cogstate; Eisai Inc.; Elan Pharmaceuticals, Inc.; Eli Lilly and Company; EuroImmun; F. Hoffmann-La Roche Ltd and its affiliated company Genentech, Inc.; Fujirebio; GE Healthcare; IXICO Ltd.; Janssen Alzheimer Immunotherapy Research & Development, LLC.; Johnson & Johnson Pharmaceutical Research & Development LLC.; Lumosity; Lundbeck; Merck & Co., Inc.; Meso Scale Diagnostics, LLC.; NeuroRx Research; Neurotrack Technologies; Novartis Pharmaceuticals Corporation; Pfizer Inc.; Piramal Imaging; Servier; Takeda Pharmaceutical Company; and Transition Therapeutics. The Canadian Institutes of Health Research is providing funds to support ADNI clinical sites in Canada. Private sector contributions are facilitated by the Foundation for the National Institutes of Health (www.fnih.org). The grantee organisation is the Northern California Institute for Research and Education, and the study is coordinated by the Alzheimer’s Therapeutic Research Institute at the University of Southern California. ADNI data are disseminated by the Laboratory for Neuro Imaging at the University of Southern California.

J-ADNI was supported by the following grants: Translational Research Promotion Project from the New Energy and Industrial Technology Development Organization of Japan; Research on Dementia, Health Labor Sciences Research Grant; Life Science Database Integration Project of Japan Science and Technology Agency; Research Association of Biotechnology (contributed by Astellas Pharma Inc., Bristol-Myers Squibb, Daiichi-Sankyo, Eisai, Eli Lilly and Company, Merck-Banyu, Mitsubishi Tanabe Pharma, Pfizer Inc., Shionogi & Co., Ltd., Sumitomo Dainippon, and Takeda Pharmaceutical Company), Japan, and a grant from an anonymous foundation.

GENIC was funded by the Estrategia de Especialización Inteligente de Canarias RIS3 de la Consejería de Economía, Industria, Comercio y Conocimiento del Gobierno de Canarias, co-funded by the Programa Operativo FEDER Canarias 2014–2020 (ProID2020010063), and Universidad Fernando Pessoa Canarias. Data used in the preparation of this article was taken from the GENIC database (Group of Neuropsychological Studies of the Canary Islands, University of La Laguna, Spain. Principal investigators: José Barroso and Daniel Ferreira, contact). The following collaborators contributed to the GENIC-database but did not participate in the analysis or writing of this report (in alphabetic order by family name): Nira Cedrés, Rut Correia, Patricia Díaz, Eloy García, Lissett González, Aída Figueroa, Nerea Figueroa, Teodoro González, Zaira González, Cathaysa Hernández, Edith Hernández, Nira Jiménez, Judith López, Cándida Lozano, Alejandra Machado, María Mata, Yaiza Molina, Antonieta Nieto, Roraima Yánez-Pérez, María Sabucedo, Elena Sirumal, Marta Suárez, Manuel Urbano, and Pedro Velasco.

## Notes

### Competing Interest Statement

The authors have declared no competing interest.

### Funding Statement

This study was funded by the Swedish Research Council (VR), the Strategic Research Programme in Neuroscience at Karolinska Institutet (StratNeuro), the Center for Innovative Medicine (CIMED), the Foundation for Geriatric Diseases at Karolinska Institutet, the regional agreement on medical training and clinical research (ALF) between Stockholm County Council and Karolinska Institutet, the Swedish Brain Foundation, the Swedish Alzheimer Foundation, the Åke Wiberg Foundation, the Olle Engkvist Byggmastare Foundation, the joint research funds of KTH Royal Institute of Technology and Stockholm County Council (HMT), Swedish Parkinson Foundation, King Gustaf V:s and Queen Victorias Foundation, David and Astrid Hagelens Foundation, Loo and Hans Ostermans Foundation, Vinnova and Digital Futures for financial support, and the NIA-supported Collaboratory on Research Definitions for reserve and resilience in cognitive ageing and dementia. The Titan X Pascal used for this research was donated by the NVIDIA Corporation.

### Author Declarations

Ethics committee/IRB of Karolinska Institutet gave ethical approval for this work.

### Summary of Updates

In this new submission, we present a deeper comparison of the developed CNN model in four different approaches, including the hold-out approach, compared to the cross-validation of the same model, including more data, and additionally, the model's performance using skull-stripped images. We show the robust performance of our developed CNN with two external datasets. We still focus on using images only having a rigid registration to the Montreal Neurological Institute - MNI - template space as input to the model. This revised version does not present the previous comparison with the Orthogonal Partial Least Square (OPLS) model and focuses on the development of our CNN model.

