## Supplementary material for "A Deep Learning Model for Brain Age Prediction Using Minimally Pre-processed T1w-images as Input"

*Corresponding authors:

### SUPPLEMENTARY MATERIAL

#### A. COHORTS DESCRIPTION

**UK Biobank** (Miller et al., 2016). This is a large, prospective multi-centre cohort study conducted across England, Scotland, and Wales. At baseline (2006-2010), about 500 000 participants aged 40-69 years were recruited, and extensive phenotypic and genotypic data were collected. Additionally, in 2014 UK biobank imaging study began with a plan to re-invite 100000 participants for brain, heart, and body imaging. The participants underwent 3T brain MRI scans. Within the MRI sample, we selected 13882 individuals without dementia, mild cognitive impairment (MCI), other neurological and psychiatric conditions, and good-to-excellent self-reported health.

**GENIC** (Ferreira et al., 2015b). This is a community-based longitudinal cohort of adults aged 29 to 85 years at baseline, who were recruited through advertisements in local schools, primary care health centres, and relatives and acquaintances of the research staff of the University of La Laguna, Tenerife, Canary Islands, Spain. We selected 299 participants without dementia, MCI, neurological, psychiatric, or other systemic diseases, as well as no evidence of pathological findings in the 3T brain MRIs (e.g., stroke, tumours, hippocampal sclerosis) or a history of substance abuse.

**ADNI** (Jack et al., 2008). Data used to prepare this article were obtained from the Alzheimer's Disease Neuroimaging Initiative (ADNI) database (adni.loni.usc.edu). The ADNI was launched in 2003 as a public-private partnership, led by Principal Investigator Michael W. Weiner, MD. The primary goal of ADNI has been to test whether serial MRI, positron emission tomography, other biological markers, and clinical and neuropsychological assessment can be combined to measure the progression of MCI and early AD. This longitudinal multi-centre study includes more than 2000 individuals in all levels of cognition (from unimpaired to cognitively impaired – including mild stages and dementia), aged 55-90 years, with available 1.5/3T brain MRIs, followed up over time. Within the available sample, we selected 1489 MRIs of 443 cognitively unimpaired individuals.

**AIBL** (Rowe et al., 2010). Data was collected by the AIBL study group. AIBL study methodology has been reported previously (Ellis et al., 2009). This is a longitudinal multi-centre study including 1100 Australian individuals that are cognitively healthy, MCI, or AD dementia, aged ≥60 years with available 1.5/3T brain MRIs. From this cohort, we selected 957 MRIs from 491 cognitively healthy individuals.

**AddNeuroMed** (Birkenbihl et al., 2021; Simmons et al., 2011). AddNeuroMed is part of the InnoMed European Union PF6 programme, designed to develop and validate novel surrogate markers in AD and includes MRI data with other biomarkers and clinical information. A total of 88 cognitively unimpaired individuals (44 females, 44 males), with 149 MRIs, with an average age of 75.1 yrs. (53.0 – 88.9 yrs.), with 1.5T brain MRIs, were included in the external dataset for validation in this study.

**JADNI** (Iwatsubo et al., 2018). Data used in the preparation of this article were obtained from the Japanese Alzheimer's Disease Neuroimaging Initiative (J-ADNI) database deposited in the National Bioscience Database Center Human Database, Japan (Research ID: hum0043.v1, 2016). The J-ADNI was launched in 2007 as a public-private partnership, led by Principal Investigator Takeshi Iwatsubo, MD. The primary goal of J-ADNI has been to test whether serial magnetic resonance imaging (MRI), positron emission tomography (PET), other biological markers, and clinical and neuropsychological assessment can be combined to measure the progression of late mild cognitive impairment (MCI) and mild Alzheimer's disease (AD) in the Japanese population. This is a large-scale observational study based on the ADNI cohort in a Japanese population and includes neuroimaging, neuropsychological and biomarker data. From this dataset, we selected 86 individuals (50 female, 36 male), with 413 MRIs, following the inclusion and exclusion criteria mentioned in section **Error! Reference source not found.**, with an average age of 68.5 yrs. (60 – 86 yrs.), and with a 1.5T and 3T (385 and 28 MRIs, respectively) brain MRI.

### B. ICD-9 AND ICD-10 CODES

Table B-1. List of ICD-10 and 9 codes of chronic diseases used as exclusion criteria in the UK Biobank. (ICD10: https://icd.who.int/browse10/2010/en#; ICD9 https://icd.codes/icd9cm).

| | ICD-10 codes | ICD-9 codes |
|---|---|---|
| **Dementia (in AD, vascular, in other diseases classified elsewhere, unspecified, delirium superimposed on dementia, AD, other neurodegenerative disorders)** | F00-F03, F051 | 290; 293-294 |
| **Cognitive impairment** | G30-G32 | |
| **Epilepsy** | G40-G41; G405 | |
| **Multiple sclerosis** | G35 | 340 + 348-349 (Other conditions of the brain) |
| **Parkinson and parkinsonism** | G20-G23 | 332 |
| **Cerebrovascular disease:** | | |
| Vascular syndromes of the brain in cerebrovascular diseases | G46 | |
| Subarachnoid haemorrhage | I60 | 430 |
| Intracerebral haemorrhage | I61 | 431 |
| Other nontraumatic intracranial haemorrhages | I62 | 432 |
| Cerebral infarction | I63 | |
| Stroke, not specified as haemorrhage or infarction | I64 | |
| Other cerebrovascular diseases | I67 | |
| Sequelae of cerebrovascular disease | I69 | 438 |
| **Schizophrenia and delusional disorders** | F20; F22; F24; F25; F28 | 295; 297 |
| **Psychiatric, mood and behavioural diseases:** | | 316 (other psychiatric factors associated with disease) |
| Manic episode | F30 | 296; 298 (incl. all below) |
| Bipolar affective disorder | F31 | |
| Recurrent depressive disorder | F33 | |
| Persistent mood [affective] disorders | F34 | |
| Other mood [affective] disorders | F38 | |
| Unspecified mood [affective] disorder | F39 | |
| Mixed anxiety and depressive disorder | F412 | |
| Dissociative [conversion] disorders | F44 | |
| Organic amnesic syndrome, not induced by alcohol and other psychoactive substances | F04 | |
| Other mental disorders due to brain damage and dysfunction and physical disease | F06 | |
| Personality and behavioural disorders due to brain disease, damage and dysfunction | F07 | |
| Unspecified organic or symptomatic mental disorder | F09 | |
| Mental and behavioural disorders due to the use of alcohol | F10 | 291; 303 (Alcohol-induced mental disorders) |
| Mental and behavioural disorders due to the use of opioids | F11 | 292; 304 (Drug-induced mental disorders) |
| Mental and behavioural disorders due to the use of cannabinoids | F12 | |
| Mental and behavioural disorders due to the use of sedatives/hypnotics | F13 | |

| | | |
|---|---|---|
| Mental and behavioural disorders due to the use of cocaine | F14 | |
| Mental and behavioural disorders due to the use of stimulants, including caffeine | F15 | |
| Mental and behavioural disorders due to the use of the hallucinogen | F16 | |
| Mental and behavioural disorders due to the use of tobacco | F17 | |
| Mental and behavioural disorders due to the use of volatile solvents | F18 | |
| Mental and behavioural disorders due to multiple drug use and use of other psychoactive substances | F19 | |
| Personality disorders | F60-F63; F68 | 301 |
| **Mental retardation** | F70-F73; F78-F79 | 317-319 (Intellectual disabilities) |
| **Developmental disorders** | F80-F84; F88-F89 | 299; 315 |
| **Mental disorder, not otherwise specified** | F99 | |

NOTE: The diseases above are chronic and known to severely affect the brain and/or cognition. Transient Ischemic Attacks (ICD-10 code G45) have not been excluded. We did not exclude cognitive impairment based on the cognitive test because only 2 tests have more than 50% of complete data in v2 (close to MRI), 399 and 20016 (fluid).

### C. CHRONOLOGICAL AGE DISTRIBUTION OF USED DATASETS IN CNN1

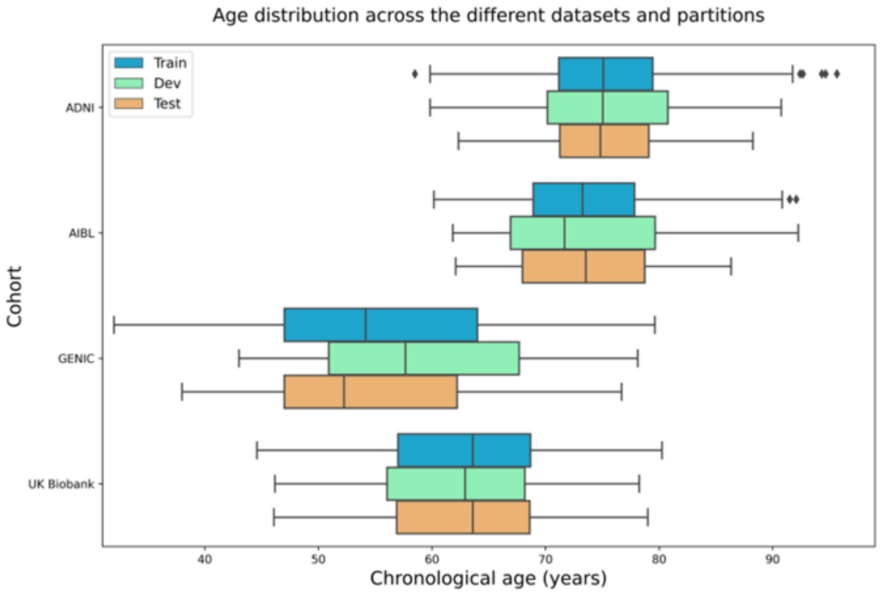


Figure C-1. Data distribution in the train, development, and test sets for the ADNI, AIBLE, Genic, and UK Biobank cohorts.

### D. MAE PER COHORT

Table D-1 presents the normalized MAE (where nMAE = (MAE/standard deviation of the cohort's population age)) of the UK Biobank.

Table D-1. Normalized MAE for each cohort for each of the trained models.

| | **Normalized MAE per Cohort** | | | | | |
|---|---|---|---|---|---|---|
| **Model** | AddNeuroMed | ADNI | AIBL | GENIC | J-ADNI | UK Biobank |
| CNN1 | 0.66 | 0.41 | 0.44 | 0.39 | 1.04 | 0.37 |
| CNN2 | 0.60 | 0.39 | 0.41 | 0.36 | 0.98 | 0.35 |
| CNN3 | 0.50 | 0.39 | 0.41 | 0.37 | 0.42 | 0.36 |
| CNN4 | 0.60 | 0.48 | 0.51 | 0.41 | 0.77 | 0.40 |

Below, in Table D-2, are the coefficient of determination ($R^2$) of the predictions for each cohort in the different trained CNNs.

Table D-2. The coefficient of determination for each cohort for each of the trained models.

| | **$R^2$ per Cohort** | | | | | |
|---|---|---|---|---|---|---|
| **Model** | AddNeuroMed | ADNI | AIBL | GENIC | J-ADNI | UK Biobank |
| CNN1 | 0.63 | 0.83 | 0.82 | 0.87 | 0.55 | 0.88 |
| CNN2 | 0.67 | 0.86 | 0.84 | 0.88 | 0.59 | 0.89 |
| CNN3 | 0.77 | 0.86 | 0.84 | 0.88 | 0.85 | 0.89 |
| CNN4 | 0.71 | 0.80 | 0.78 | 0.87 | 0.64 | 0.85 |

### E. RELEVANT BRAIN REGIONS FOR AGING PREDICTION

The relevant brain regions for aging prediction were calculated based on the salience maps, constructed using the SmoothGrad approach, with a kernel smoothing of 0.15. Salience maps of all individuals were calculated individually and averaged through the whole sample. Figure E-1 presents the averaged maps of the 1% percentiles of the highest absolute values of the salience maps for CNN1, 2, 3 and 4, respectively.

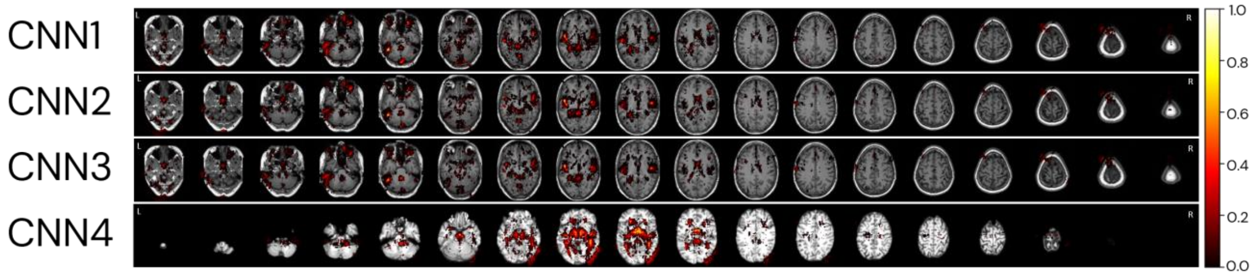


Figure E-1. Averaged salience map of the trained models.

### F. BRAIN AGE DIFFERENCES FOR SELECTED INDIVIDUALS

Calculated brain age differences for CNN1, 2, 3 and 4, for the same individuals, used to create Figure 10, in the main manuscript. The chronological age (CA), and the brain age difference to the chronological age (GAP) of the individuals are presented in Table F-1.

Table F-1. Calculated brain age difference between chronological and predicted brain age for each model for the randomly selected individuals to plot Figure 10 in the manuscript.

| Cohort | GAP considered | CA | GAP_CNN1 | GAP_CNN2 | GAP_CNN3 | GAP_CNN4 |
|---|---|---|---|---|---|---|
| UK Biobank | -8 | 65.25 | -8.9 | -8.0 | -8.5 | -8.0 |
| UK Biobank | -7 | 65.42 | -7.2 | -7.5 | -8.0 | -4.3 |
| UK Biobank | -6 | 64.83 | -6.8 | -5.6 | -5.5 | -1.8 |
| UK Biobank | -5 | 65.42 | -5.7 | -4.6 | -5.0 | 2.0 |
| UK Biobank | -4 | 65.50 | -4.8 | -4.4 | -4.8 | -0.5 |
| UK Biobank | -3 | 65.33 | -3.7 | -4.0 | -4.2 | -0.9 |
| UK Biobank | -2 | 65.08 | -2.2 | -0.7 | -0.4 | 4.7 |
| UK Biobank | -1 | 65.33 | -1.6 | -2.0 | -1.8 | 1.4 |
| UK Biobank | 0 | 64.25 | -0.5 | -0.6 | -0.8 | -3.0 |
| UK Biobank | 0 | 65.33 | 0.6 | 0.5 | 0.3 | 0.7 |
| UK Biobank | 1 | 64.58 | 1.0 | -0.1 | -0.1 | -6.0 |
| UK Biobank | 2 | 65.08 | 2.1 | 0.7 | 0.3 | -0.3 |
| AIBL | 3 | 65.58 | 3.1 | 3.0 | 3.3 | 5.2 |
| UK Biobank | 4 | 65.50 | 4.3 | 4.2 | 4.7 | 6.7 |
| UK Biobank | 5 | 65.83 | 5.1 | 4.8 | 4.8 | 3.6 |
| UK Biobank | 6 | 65.17 | 6.0 | 5.5 | 5.0 | 1.3 |
| AIBL | 7 | 65.33 | 6.7 | 3.0 | 3.3 | -0.7 |
| ADNI | 8 | 65.17 | 8.2 | 5.6 | 5.9 | 3.4 |

### G. INDIVIDUALS IN EACH AGE GROUP FOR THE DIFFERENTIAL ANALYSIS IN NEUROIMAGING STUDIES

For the generated image of group comparison of age and predicted brain age with a younger group, individuals were grouped based on chronological and predicted brain age. The groups are referent to the age±1 year. Table G-1shows the number of individuals in each age group used in the analysis.

Table G-1. Number of individuals in each age group for the differential analysis on using chronological or predicted brain age in neuroimaging studies.

| **Age group** | **Chronological age** | **Predicted brain age** |
|---|---|---|
| 55 | 904 | 977 |
| 60 | 1131 | 1082 |
| 65 | 1474 | 1559 |
| 70 | 1542 | 1714 |
| 75 | 797 | 493 |
| 80 | 216 | 255 |
